# Evaluating the Influence of Mood and Stress on Glycemic Variability in T1DM Patients using Glucose Monitoring Sensors and Pools

**DOI:** 10.1101/2020.09.03.20186940

**Authors:** Sergio Contador, Marta Botella, Aranzazu Aramendi, Remedios Martínez, Esther Maqueda, Oscar Garnica, J. Manuel Velasco, Juan Lanchares, J. Ignacio Hidalgo

## Abstract

**Objective:** Assess in a sample of patients with type 1 diabetes mellitus whether mood and stress influence blood glucose levels and variability.

**Material and Methods:** Continuous glucose monitoring was performed on 10 patients with type 1 diabetes, where interstitial glucose values were recorded every 15 minutes. A daily survey was conducted through Google Forms, collecting information on mood and stress. The day was divided into 6 slots of 4-hour each, asking the patient to assess each slot in relation to mood (sad, normal or happy) and stress (calm, normal or nervous). Different measures of glycemic control (arithmetic mean and percentage of time below/above the target range) and variability (standard deviation, percentage coefficient of variation, mean amplitude of glycemic excursions and mean of daily differences) were calculated to relate the mood and stress perceived by patients with blood glucose levels and glycemic variability. A hypothesis test was carried out to quantitatively compare the data groups of the different measures using the Student’s t-test.

**Results:** Statistically significant differences (p-value < 0.05) were found between different levels of stress. In general, average glucose and variability decrease when the patient is calm. There are statistically significant differences (p-value < 0.05) between different levels of mood. Variability increases when the mood changes from sad to happy. However, the patient’s average glucose decreases as the mood improves.

**Conclusions:** Variations in mood and stress significantly influence blood glucose levels, and glycemic variability in the patients analyzed with type 1 diabetes mellitus. Therefore, they are factors to consider for improving glycemic control. The mean of daily differences does not seem to be a good indicator for variability.

## 1 Background

The prevalence of depression and psychosocial stress is higher in patients with Type 1 Diabetes Mellitus (T1DM) [1]. The presence of emotional disorders has been associated with poorer long-term glycemic control [2]. The impact of daily mood swings and stress on short-term glycemic control and glycemic variability has been little studied, and to date no study with continuous glucose monitoring has been reported. High glycemic variability increases the risk of hypoglycemia, hinders metabolic control, and may be associated with an increase in chronic complications.

Stress is not only related to high blood glucose levels, but there are people in whom stress causes low blood glucose levels [3]. With stress, they lose their appetite and increase their risk of life-threatening hypoglycemia. In some cases, it could lead them to present what is called chronic fatigue [4]. Significant associations between stress and metabolic control have been found in several studies, through the HbA1c level in patients with diabetes [5, 6, 7, 8]. Other studies use the The Diabetes Distress Scale [9], where stress was found to be directly and statistically significantly correlated with HbA1c level [10].

This work aims to study whether mood changes and stress influence blood glucose levels in the short term, relating mood and stress perceived by patients with blood glucose levels and glycemic variability. The study is performed by analyzing two glycemic control metrics (arithmetic mean and percentage of time below/above the target range) and four glycemic variability metrics (standard deviation, percentage coefficient of variation, mean amplitude of glycemic excursions and mean of daily differences).

The rest of the article is organized as follows. Section 2 describes the data and the measures used for glycemic control and the calculation of variability. The results obtained are shown in section 3. The conclusions are presented in section 4.

## 2 Material and Methods

### 2.1 Patients

Continuous Glucose Monitoring (CGM) was performed using Free style Libre sensors from Abbott, on 10 patients with T1DM where interstitial glucose values were recorded every 15 minutes. A daily survey was developed through Google Forms, collecting information on mood and stress. The form is available through an online link (https://forms.gle/ehcoo6hGrbKouscMA). The patients filled out the form daily, recording in each survey the data relative to the previous day. In addition, it was indicated that the form must be completed for each day the patient participated in the study and the patient was reminded that it was essential that the information was as accurate as possible. It was also stated that in the case in which the patient forgot to write down a value and did not remember it, it would be preferable to tag as *I don’t remember* this slot.

The survey asked about the patient’s feeling regarding the mood and stress. For this, the day was divided into 6 slots of 4-hour each starting from 00:00h, and the patient was asked to assess each slot in relation to the mood (sad, normal or happy) and stress (calm, normal or nervous). In addition, data were collected on the quality and duration of nighttime sleep and any naps. The current form includes other options, such as anger, incorporated at the suggestion of the patients.

Table 1 shows the number of 4-hour slots of data per patient for each of the categories of the variables of the survey stress and mood. The number of days with valid data per patient is also displayed. Not every day has the same number of slots, nor do all the slots have the same number of glucose values. Most days (96.6 [%]) have all the slots completed (6 slots per day) with all glucose values (97.4 [%]) filled (16 values per slot).

**Table 1:** Number of days with valid measurements and number of 4-hour slots valid for each category of stress and mood.

| Patient | P1 | P2 | P3 | P4 | P5 | P6 | P7 | P8 | P9 | P10 | Total |
| --- | --- | --- | --- | --- | --- | --- | --- | --- | --- | --- | --- |
| Days | 50 | 13 | 16 | 11 | 4 | 50 | 14 | 8 | 16 | 10 | 192 |
| 4-hour slots with valid data and stress survey per patient |  |  |  |  |  |  |  |  |  |  |  |
| Calm | 0 | 0 | 1 | 0 | 8 | 0 | 0 | 0 | 5 | 0 | 14 |
| Normal | 287 | 55 | 68 | 51 | 7 | 44 | 61 | 39 | 74 | 46 | 732 |
| Nervous | 13 | 21 | 20 | 15 | 3 | 26 | 21 | 4 | 17 | 11 | 151 |
| 4-hour slots with valid data and mood survey per patient |  |  |  |  |  |  |  |  |  |  |  |
| Sad | 0 | 6 | 5 | 5 | 0 | 1 | 4 | 0 | 11 | 4 | 36 |
| Normal | 266 | 72 | 62 | 61 | 16 | 36 | 77 | 30 | 79 | 46 | 745 |
| Happy | 34 | 0 | 22 | 0 | 2 | 0 | 1 | 13 | 6 | 7 | 85 |

When collecting data with CGM sensors for long periods of time, it is common to find some data that has not been recorded. To solve this problem, we performed an attribution of the values using segmental interpolation (splines) of degree 3 [11], where the maximum number of consecutive values to be filled is 4 (maximum correction of 1 hour).

Table 2 shows the clinical data of each patient. 60 [%] of the patients are women. The mean HbA1c value is 7.0*±*0.8 [%], with a mean weight of 62.5*±*10.2 [Kg], a mean age of 29.1*±*10.9 [years], a mean height of 163.7*±*11.2 [cm] and a mean DX time of 16.5*±*9.0 [years]. Half of the patients undergo insulin treatment using Multiple Dose of Insulin (MDI), and the other half via Continuous Infusion of Subcutaneous Insulin (ICIS).

**Table 2:** Clinical characterization of each patient. Insulin treatment is performed by Multiple Dose of Insulin (MDI) or by Continuous Subcutaneous Insulin Infusion (ICIS).

| Measure | P1 | P2 | P3 | P4 | P5 | P6 | P7 | P8 | P9 | P10 |
| --- | --- | --- | --- | --- | --- | --- | --- | --- | --- | --- |
| Gender | Woman | Man | Man | Woman | Woman | Woman | Woman | Man | Man | Woman |
| HbA1c [%] | 6.9 | 7.5 | 7.0 | 6.5 | 7.6 | 5.4 | 8.3 | 7.1 | 6.7 | 7.2 |
| Age [years] | 10 | 25 | 43 | 45 | 27 | 26 | 33 | 18 | 26 | 38 |
| DX time [years] | 2 | 14 | 12 | 35 | 16 | 20 | 16 | 8 | 24 | 18 |
| Weight [kg] | 39.0 | 60.0 | 63.0 | 70.0 | 58.6 | 67.0 | 57.2 | 69.7 | 77.0 | 63.4 |
| Height [cm] | 140 | 159 | 165 | 169 | 159 | 164 | 167 | 168 | 185 | 161 |
| ICIS/MDI | MDI | MDI | MDI | ICIS | ICIS | ICIS | ICIS | MDI | MDI | ICIS |

The inclusion criteria of the patients in the study were as follows:

- Patients with T1DM of at least 1 year since DX.
- Absence of diagnosis of major psychiatric disorder.
- No serious breakthrough disease in the last 6 months.

The study was approved by the Ethics Committee of the Hospital Príncipe de Asturias and all patients signed a prior informed consent.

### 2.2 Methodology

In addition to the HbA1c level, there are different glycemic control measures such as the arithmetic mean or the percentage of time below/above the target range [12], among others, and numerous measures that have been developed to assess glycemic variability. The following measures have been used in this study as glycemic control parameters, as described in the recommendations of the American Diabetes Association [13]:

- Average blood glucose (MEAN).
- Percentage of time in hypoglycemia level 1 (hp1) (< 70 [mg/dl]) and level 2 (hp2) ([54,70) [mg/dl]).
- Percentage of time in range (Range) ([70,180] [mg/dl]).
- Percentage of time in hyperglycemia of level 1 (HP1) (*>* 180 [mg/dl]) and level 2 (HP2) (*>* 250 [mg/dl]). As variability parameters, the following were analyzed:
- Mean standard deviation (SD) evaluated every 4 hours (SD4h) and 24 hours (SD24h).
- Percentage variation coefficient (CV) evaluated every 4 hours (CV4h) and 24 hours (CV24h).
- Average amplitude of the glycemic excursions (MAGE) evaluated every 4 hours (MAGE4h) and 24 hours (MAGE24h).
- Average of daily differences (MODD).

The mood and stress perceived by the patients were correlated with blood glucose levels and glycemic variability. To do this, a hypothesis test was carried out to quantitatively compare the data groups of the different measures of variability. We applied the Student’s t-test (parametric test) using a confidence level of 95 [%] (*α* = 0.05).

## 3 Results

Table 3 shows the different measures of glycemic variability and glycemic averages along with their standard deviations for the 10 patients (P1 to P10). Figure 1 shows the interquartile ranges of glucose values along with their mean values, and the time in range of each patient.

**Table 3:** Averages of the different measures of glycemic variability and glycemic averages together with their standard deviations for the 10 patients (P1 to P10).

| Measure | P1 | P2 | P3 | P4 | P5 | P6 | P7 | P8 | P9 | P10 |
| --- | --- | --- | --- | --- | --- | --- | --- | --- | --- | --- |
| MEAN [mg/dl] | 145.46 | 158.58 | 142.00 | 199.46 | 139.66 | 119.69 | 143.84 | 156.14 | 136.53 | 145.07 |
| SD [mg/dl] | 53.41 | 69.13 | 54.75 | 61.90 | 63.76 | 45.39 | 70.67 | 58.26 | 56.34 | 58.21 |
| hp1 [%] | 3.51 | 5.37 | 4.05 | 0.57 | 3.86 | 10.22 | 6.42 | 3.44 | 6.31 | 6.40 |
| hp2 [%] | 0.61 | 0.96 | 1.85 | 0.00 | 16.14 | 4.17 | 11.07 | 0.00 | 3.02 | 0.99 |
| Rango [%] | 69.97 | 60.66 | 69.87 | 37.29 | 51.58 | 75.16 | 51.39 | 62.93 | 72.67 | 65.05 |
| HP1 [%] | 25.91 | 33.01 | 24.24 | 62.14 | 28.42 | 10.45 | 31.11 | 33.63 | 18.00 | 27.56 |
| HP2 [%] | 3.95 | 11.62 | 2.99 | 17.88 | 2.81 | 0.48 | 7.20 | 6.58 | 3.28 | 3.42 |
| MODD [mg/dl] | 56.87±29.73 | 78.47±48.15 | 62.55±33.09 | 64.75±40.77 | 30.31±12.91 | 48.65±24.96 | 63.50±34.29 | 60.63±27.23 | 56.49±24.04 | 69.31±25.02 |
| SD4h [mg/dl] | 30.14±15.88 | 33.25±16.67 | 25.36±15.00 | 27.23±13.57 | 24.68±18.24 | 25.40±13.40 | 35.11±22.66 | 38.77±16.08 | 27.36±16.98 | 31.44±17.56 |
| SD24h [mg/dl] | 47.92±11.43 | 54.73±16.45 | 49.28±11.39 | 53.10±7.85 | 41.16±24.57 | 40.90±9.17 | 61.45±16.34 | 54.86±9.75 | 46.78±15.63 | 51.52±14.17 |
| MAGE4h [mg/dl] | 19.60±10.51 | 18.57±9.34 | 11.70±7.32 | 13.71±7.46 | 11.32±8.33 | 14.82±8.78 | 13.10±10.05 | 19.93±13.83 | 11.70±7.29 | 19.09±10.10 |
| MAGE24h [mg/dl] | 31.51±7.75 | 28.60±10.66 | 21.74±9.17 | 27.16±7.94 | 12.19±10.48 | 26.03±8.75 | 21.31±10.44 | 37.33±6.85 | 23.12±6.89 | 31.28±8.05 |
| CV4h [%] | 21.50±10.51 | 22.51±11.60 | 18.80±10.29 | 14.44±7.99 | 18.63±14.20 | 22.10±10.96 | 26.66±15.57 | 25.72±11.04 | 21.09±12.31 | 22.81±11.42 |
| CV24h [%] | 33.04±6.80 | 35.12±8.48 | 34.75±6.55 | 27.06±4.63 | 27.59±13.10 | 34.69±8.55 | 42.95±7.05 | 35.06±4.76 | 34.64±9.67 | 35.97±8.79 |

**Figure 1:**
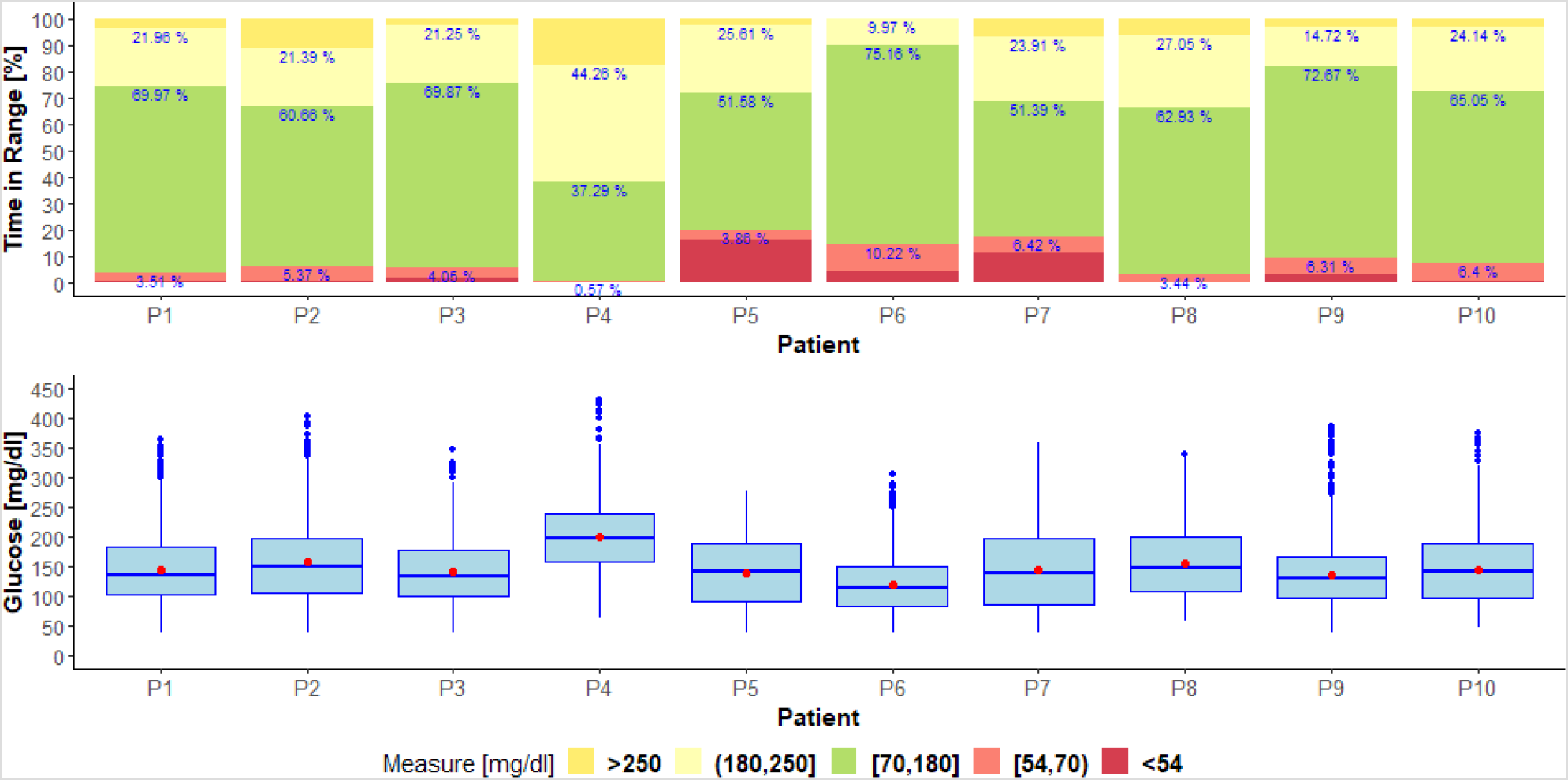
The figure above shows the percentage of time the patient has a very low glucose level (< 54 [mg/dl]), low ([54,70) [mg/dl]), in range ([70,180] [mg/dl]), high ((180,250] [mg/dl]), and very high (*>* 250 [mg/dl]). The figure below shows the interquartile ranges of glucose. The mean value is represents with a red dot.

MEAN, SD, hp1, hp2, Range, HP1 and HP2 measurements were calculated using all glucose values. The SD24h, MAGE24h and CV24h measurements were calculated using the mean glucose values in 24-hour slots (one full day). The MODD, SD4h, MAGE4h and CV4h measurements were calculated using the mean of glucose values in 4-hour slots.

To correlate the stress and mood with blood glucose levels, the combined categories of the two variables have been taken into account. Tables 4 and 5 show the results obtained for the stress. Tables 6 and 7 for mood. The tables show the averages of the different measures of glycemic variability and glycemic averages for the different categories of the variables, and the p-values obtained by hypothesis testing for the combined categories.

**Table 4:** Averages of the different measures of glycemic variability and glycemic averages for the different categories of the stress variable.

| Average | Calm | Normal | Nervous |
| --- | --- | --- | --- |
| MEAN | 126.56 | 147.96 | 146.13 |
| SD | 23.55 | 29.31 | 32.95 |
| CV | 20.82 | 20.94 | 24.39 |
| MAGE | 10.97 | 16.27 | 17.27 |
| MODD | 66.59 | 60.19 | 61.67 |

**Table 5:**
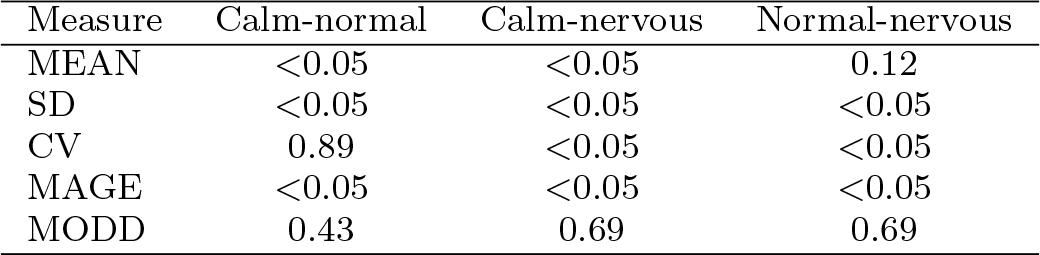
P-values obtained by hypothesis testing for the different measures of glycemic variability and glycemic averages for the different categories of the stress variable.

| Measure | Calm-normal | Calm-nervous | Normal-nervous |
| --- | --- | --- | --- |
| MEAN | $<0.05$ | $<0.05$ | 0.12 |
| SD | $<0.05$ | $<0.05$ | $<0.05$ |
| CV | 0.89 | $<0.05$ | $<0.05$ |
| MAGE | $<0.05$ | $<0.05$ | $<0.05$ |
| MODD | 0.43 | 0.69 | 0.69 |

**Table 6:** Averages of the different measures of glycemic variability and glycemic averages for the different categories of the mood variable.

| Average | Sad | Normal | Happy |
| --- | --- | --- | --- |
| MEAN | 151.88 | 148.00 | 146.52 |
| SD | 34.39 | 29.04 | 36.75 |
| CV | 24.53 | 20.99 | 25.14 |
| MAGE | 15.43 | 16.03 | 20.41 |
| MODD | 58.75 | 61.05 | 59.95 |

**Table 7:** P-values obtained by hypothesis testing for the different measures of glycemic variability and glycemic averages for the different categories of the mood variable.

| Measure | Sad-normal | Sad-happy | Normal-happy |
| --- | --- | --- | --- |
| MEAN | 0.15 | 0.06 | 0.18 |
| SD | <0.05 | <0.05 | <0.05 |
| CV | <0.05 | 0.33 | <0.05 |
| MAGE | 0.10 | <0.05 | <0.05 |
| MODD | 0.79 | 0.89 | 0.77 |

Here follows a study of the results obtained for the stress. Statistically significant differences have been found (p-value < 0.05):

- In the MEAN measure, where the mean increases when the stress increases (calm-normal 126.56 vs 147.96 [mg/dl], calm-nervous 126.56 vs 146.13 [mg/dl]).
- In the SD measure, where the variability (in all categories) increases when the stress increases.
- In the CV measure, where the variability (in all categories) increases when the stress increases.
- In the MAGE measure, where the variability (in all categories) increases when the stress increases.

No statistically significant differences (p-value < 0.05) were found in the MODD measure.

Here follows a study of the results obtained for mood. No statistically significant differences were found (p-value < 0.05):

- In the MEAN measure, however, the patient’s mean glucose (in all categories) decreases as mood improves.
- Applying MODD.

Statistically significant differences have been found (p-value < 0.05):

- In the SD measure, where the variability increases when the mood changes from sad to happy.
- In the CV measure, where the variability increases when the mood changes from sad to happy.
- In the MAGE measure, where the variability (in all categories) increases when mood improves.

## 4 Conclusions

Variations in mood and stress significantly influence blood glucose levels in patients with T1DM according to the measurements analyzed in this study.

In general, stress increases the patient’s mean and glycemic variability. This trend has been demonstrated in all the categories studied. The mood results have not been as conclusive. The glycemic mean decreases as the patient’s mood improves. However, glycemic variability, in general, increases when there is a change in mood, whether the patient is sad or happy, although there are discordant results regarding the different measures of variability.

The MODD measure is not shown as a good indicator of variability since it is the only measure that has not found significant differences in any compared group.

Stress and mood affect the blood glucose levels of patients and are, therefore, factors to consider for improving glycemic control.

## Data Availability

Data of the patients is not publicly available. It could be available on demand and after signing a Non Discluser Agreement.

## Acknowledgements

This work has been funded by:

- Fundación Eugenio Rodríguez Pascual 2019-2020.
- Ministerio de Economía y Competitividad under grant TIN2014-54806-R.
- Ministerio de Ciencia, Innovación y Universidades under grant RTI2018-095180-B-I00.
- Comunidad de Madrid under grants B2017/BMD3773 (GenObIA-CM) and Y2018/NMT-4668 (Micro-Stress-MAP-CM).
- European Union through structural and FEDER Funds.

## References

[1] Ryan J. Anderson, Kenneth E. Freedland, Ray E. Clouse, and Patrick J. Lustman. The prevalence of comorbid depression in adults with diabetes. Diabetes Care, 24(6):1069–1078, 2001.

[2] Patrick J. Lustman and Ray E. Clouse. Depression in diabetic patients: The relationship between mood and glycemic control. Journal of Diabetes and its Complications, 19(2):113 – 122, 2005.

[3] E. Carrasco et al. Manual para educadores en diabetes mellitus, programa de educación en diabetes. Organización Panamericana de la Salud, september 2007.

[4] R. Surwit. Diabetes tipo 2 y estrés. Diabetes Voice, 47(4):38–40, 2002.

[5] Marcel C. Adriaanse, Frans Pouwer, Jacqueline M. Dekker, Giel Nijpels, Coen D. Stehouwer, Robert J. Heine, and Frank J. Snoek. Diabetes-related symptom distress in association with glucose metabolism and comorbidity. Diabetes Care, 31(12):2268–2270, 2008

[6] L. Fisher, M. M. Skaff, J. T. Mullan, P. Arean, R. Glasgow, and U. Masharani. A longitudinal study of affective and anxiety disorders, depressive affect and diabetes distress in adults with type 2 diabetes. Diabetic Medicine, 25(9):1096–1101, 2008.

[7] Lawrence Fisher, Joseph T. Mullan, Marilyn M. Skaff, Russell E. Glasgow, Patricia Arean, and Danielle Hessler. Predicting diabetes distress in patients with type 2 diabetes: A longitudinal study. Diabetic medicine: a journal of the British Diabetic Association, 26:622–7, 07 2009

[8] Manuel Ortiz, Eugenia Ortiz, Alejandro Gatica, and Daniela Gómez. Factores psicosociales asociados a la adherencia al tratamiento de la diabetes mellitus tipo 2. Terapia psicológica, 29:5 – 11, 07 2011.

[9] William H. Polonsky, Lawrence Fisher, Jay Earles, R. James Dudl, Joel Lees, Joseph Mullan, and Richard A. Jackson. Assessing psychosocial distress in diabetes. Diabetes Care, 28(3):626–631, 2005

[10] Bijal Shah, Gireesh Gupchup, Matthew Borrego, Dennis Raisch, and Katherine Knapp. Depressive symptoms in patients with type 2 diabetes mellitus: Do stress and coping matter? Stress and health: journal of the International Society for the Investigation of Stress, 28:111–22, 04 2012.

[11] Randall Dougherty, Alan Edelman, and James Hyman. Nonnegativity, monotonicity, or convexity-preserving cubic and quintic hermite interpolation. Mathematics of Computation, 52:471–494, 4 1989.

[12] Gabor Marics, Zsofia Lendvai, Csaba Lodi, Levente Koncz, David Zakarias, Gyorgy Schuster, Borbala Mikos, Csaba Hermann, Attila J. Szabo, and Peter Toth-Heyn. Evaluation of an open access software for calculating glucose variability parameters of a continuous glucose monitoring system applied at pediatric intensive care unit. BioMedical Engineering OnLine, 14(1):37, April 2015

[13] Thomas Danne, Revital Nimri, Tadej Battelino, Richard M. Bergenstal, Kelly L. Close, J. Hans DeVries, Satish Garg, Lutz Heinemann, Irl Hirsch, Stephanie A. Amiel, Roy Beck, Emanuele Bosi, Bruce Buckingham, Claudio Cobelli, Eyal Dassau, Francis J. Doyle, Simon Heller, Roman Hovorka, Weiping Jia, Tim Jones, Olga Kordonouri, Boris Kovatchev, Aaron Kowalski, Lori Laffel, David Maahs, Helen R. Murphy, Kirsten Nørgaard, Christopher G. Parkin, Eric Renard, Banshi Saboo, Mauro Scharf, William V. Tamborlane, Stuart A. Weinzimer, and Moshe Phillip. International consensus on use of continuous glucose monitoring. Diabetes Care, 40(12):1631–1640, 2017

